# Assessment of the Reliability, Responsiveness, and Meaningfulness of the Scale for the Assessment and Rating of Ataxia (SARA) for Lysosomal Storage Disorders

**DOI:** 10.1101/2024.07.19.24310680

**Authors:** Julien Park, Tatiana Bremova-Ertl, Marion Brands, Tomas Foltan, Matthias Gautschi, Paul Gissen, Andreas Hahn, Simon Jones, Laila Arash-Kaps, Miriam Kolnikova, Marc Patterson, Susan Perlman, Uma Ramaswami, Stella Reichmannová, Marianne Rohrbach, Susanne Schneider, Aasef Shaikh, Siyamini Sivananthan, Matthis Synofzik, Mark Walterfarng, Pierre Wibawa, Kyriakos Martakis, Mario Manto

**Affiliations:** Department of General Paediatrics, University of Münster, 48149 Münster, Germany; Department of Neurology, University Hospital Bern (Inselspital), Switzerland; Department of Paediatric Metabolic Disease Amsterdam University Medical Center, Amsterdam, Netherlands; Department of Pediatric Neurology, National Institute of Children’s Diseases, Comenius University in Bratislava, Bratislava, Slovak Republic; Division of Paediatric Endocrinology, Diabetology and Metabolism, Department of Paediatrics, and Institute of Clinical Chemistry, Inselspital, Bern University Hospital, University of Bern, Switzerland; NIHR Great Ormond Street Hospital Biomedical Research Centre, University College London, United Kingdom; Department of Child Neurology, Justus Liebig University, Giessen, Germany; Willink Unit, Manchester Centre for Genomic Medicine, Royal Manchester Children’s Hospital, University of Manchester, United Kingdom; SphinCS - Institute of Clinical Science in Lysosomal Storage Disorders, Hochheim, Germany; Departments of Neurology, Pediatrics and Clinical Genomics, Mayo Clinic Children’s Center, Rochester, MN, USA; Department of Neurology, University of California Los Angeles, CA, United States; Lysosomal Storage Disorder Unit, Royal Free London NHS Foundation Trust, United Kingdom; Department of Paediatrics and Inherited Metabolic Disorders, First Faculty of Medicine, Charles University and General University Hospital in Prague, Czech Republic; Division of Metabolism, University Children’s Hospital and Children’s Research Centre, Zurich, Switzerland; Department of Neurology, Ludwig Maximilians University, Munich, Germany; Department of Neurology, Case Western Reserve University School of Medicine Cleveland, OH, USA; Department of Neurodegenerative Diseases, Hertie-Institute for Clinical Brain Research and Center of Neurology, University of Tübingen, Tübingen, Germany; Department of Neuropsychiatry, The Royal Melbourne Hospital, Melbourne, Australia; Department of Pediatrics, Medical Faculty and University Hospital, University of Cologne, Cologne, Germany; Unité des Ataxies Cérébelleuses, Service de Neurologie, Médiathèque Jean Jacquy, CHU-Charleroi, 6000, Charleroi, Belgium

**Author notes:** **Correspondence to**: Dr Julien Park, Department of General Paediatrics, University of Münster, 48149 Münster, Germany.

**Keywords:** Scale for the Assessment and Rating of Ataxia, Lysosomal Storage Disorders, Clinical Outcome Assessments

## Abstract

**Objective:** To evaluate the reliability, responsiveness, and validity of the Scale for the Assessment and Rating of Ataxia (SARA) in patients with Lysosomal Storage Disorders (LSDs) who present with neurological symptoms and quantify the threshold for a clinically meaningful change.

**Methods:** We analyzed data from three clinical trial cohorts (IB1001-201, IB1001-202, IB1001-301) of patients with Niemann-Pick disease type C (NPC) and GM2 Gangliosidoses (Tay-Sachs and Sandhoff disease) comprising 122 patients and 703 visits. Reproducibility was described as retest reliability between repeat baseline visits or baseline vs. post-treatment washout visits. Responsiveness was determined in relation to the Investigator’s, Caregiver’s, and Patient’s Clinical Global Impression of Improvement (CGI-I). The CGI-I data was also used to quantify a threshold for a clinically meaningful improvement on the SARA scale. Using a qualitative methods approach, patient/caregiver interviews from the IB1001-301 trial were further used to assess a threshold of meaningful change as well as the breadth of neurological signs and symptoms captured and evaluated by the SARA scale.

**Results:** The Inter-Class Correlation (ICC) was 0.95 or greater for all three trials, indicating a high internal consistency/reliability. The mean change in SARA between repeat baseline or post-treatment washout visit assessments in all trials was -0.05, SD 1.98, i.e. minimal, indicating no significant differences, learning effects or other systematic biases. For the CGI-I responses and change in SARA scores Area Under the Curve (AUC) values were 0.82, 0.71, and 0.77 for the Investigator’s, Caregiver’s, and Patient’s CGI-I respectively, indicating strong agreement. Further qualitative analyses of the patient/caregiver interviews demonstrated a 1-point or greater change on SARA to be a clinically meaningful improvement which is directly relevant to the patient’s everyday functioning and quality of life. Changes captured by the SARA were also paralleled by improvement in a broad range of neurological signs and symptoms and beyond cerebellar ataxia.

**Conclusion:** Qualitative and quantitative data demonstrate the reliability and responsiveness of the SARA score as a valid measure of neurological signs and symptoms in LSDs. A 1-point change represents a clinically meaningful transition reflecting the gain or loss of complex function.

**Statements and Declarations:** The IB1001-201, IB1001-202, and IB1001-301 trials were funded by IntraBio Inc. No funding was received to assist with the preparation of this manuscript.

T. Bremova-Ertl received honoraria for lecturing from Sanofi and Acetlion and fees from IntraBio to serve as a blinded rater for the IB1001-201 and IB1001-202 clinical trials. P. Gissen received consulting fees from Mandos Health and is the Co-Founder and shareholder of Bloomsbury Genetic Therapies. M. Patterson is a shareholder in IntraBio and his institution has received research grants from Azafaros, Glycomine, Idorsia, Maggie’s Pearl, Takeda, and Zevra, and consulting fees (directed to his institution) from Zevra. U. Ramaswami has received research and/or investigator-initiated research grants from Amicus and Takeda and honoraria for advisory boards and lectures from Amicus, Takeda, and Sanofi. All other authors The authors have no competing interests to declare that are relevant to the content of this article.

## Introduction

### Scale for the Assessment and Rating of Ataxia (SARA)

The Scale for the Assessment and Rating of Ataxia (SARA) was initially developed to be a reliable and valid scale measuring the severity of cerebellar ataxia [1–3]. The SARA scale is composed of 8 functional domain (“item”) assessments (Gait (0-8 points), Stance (0-6 points), Sitting (0-4 points), Speech disturbance (0-6 points), Finger chase (0-4 points), Nose-finger test (0-4 points), Fast alternating hand movement (0-4 points), Heel-shin slide (0-4 points)) with total scores ranging from 0 (normal) to 40 (most severe).

The SARA underwent a rigorous validation procedure involving three large multi-center trials in spinocerebellar ataxias (SCAs) and non-spinocerebellar ataxia patients, as well as controls, which found excellent interrater reliability, test-retest reliability, and high internal consistency [2] and has undergone thorough item-response testing for multiple ataxias [4]. The SARA has also been shown to have excellent concurrent validity with other COAs, including the International Cooperative Ataxia Rating Scale (ICARS) [5], Barthel Index or with Unified Huntington’s Disease Rating Scale. Multiple studies have demonstrated that the scale reflects patient-reported symptoms and the impact of illness in cerebellar motor ataxia disorders and accurately represents patient experience [6–8]. The correlations between total SARA score and measures of daily activities and functional assessment are well-established in patients with inherited cerebellar ataxias, allowing further practical translation into the patient’s everyday life (**Table 1** provides an overview of each of the 8 SARA test items and the patient-reported activities impacted to which each test item relates to) [4, 9]. Multiple clinical studies validating the psychometric properties of the SARA scale in patients with inherited cerebellar ataxias showed an individual decrease (improvement) in the total SARA of at least 1 to 1.5 points as a clinically relevant improvement, and a decrease of 1.1 points at the group level to be clinically relevant [2].

**Table 1:** Overview of SARA test items.

| SARA Item | Test Instructions [1] | Test Description | Specific Neurological Features | Patient-reported activities impacted by specific symptom [8] |
| --- | --- | --- | --- | --- |
| **Gait** | Proband is asked (1) to walk at a safe distance parallel to a wall including a half-turn (turn around to face the opposite direction of gait) and (2) to walk in tandem (heels to toes) without support | Assessment of neurological function that measures ambulation, balance, muscle strength, coordination, and postural stability | • Ataxia<br>• Dysmetria<br>• Dystonia<br>• Dyskinesias<br>• Hypotonia<br>• Spasticity<br>• Slowing of rapid alternating movements<br>• Balance problems<br>• Muscle weakness<br>• Loss of muscle coordination | • Walking<br>◦ Walking alone, walking in crowds, walking outside on uneven surfaces, walking dog<br>• Exercise<br>• Leaving the house alone<br>• Cannot carry things as need to hold walking device<br>• Cannot perform house work<br>• Cannot travel (to and from job, to run errands, through airport, etc).<br>• Falling |
| **Stance** | Proband is asked to stand (1) in natural position, (2) with feet together in parallel (big toes touch each other) and (3) in tandem (both feet on one line, no space between heel and toe). Proband does not wear shoes, eyes are open. For each condition, three trials are allowed. Best trial is rater. | Assessment of neurological function that measures balance, muscle strength, postural stability | • Ataxia<br>• Dysmetria<br>• Dystonia<br>• Dyskinesias<br>• Hypotonia<br>• Spasticity<br>• Slowing of rapid alternating movements<br>• Balance problems<br>• Muscle weakness<br>• Loss of muscle coordination | • Standing up<br>• Standing in the shower<br>• Standing in line<br>• Cannot socialize (cannot hold drink in conversation)<br>• Cannot step on/off curb without aid<br>• Housework<br>• Playing Sports<br>• Cannot squat or reach up<br>• Falling |
| **Sitting** | Proband is asked to sit on an examination bed without support of feet, eyes open and arms outstretched to the front. | Assessment of neurological function that measures balance, muscle strength, postural stability | • Dysmetria<br>• Ataxia<br>• Dystonia<br>• Dyskinesias<br>• Hypotonia<br>• Spasticity<br>• Balance Problems<br>• Muscle Weakness<br>• Loss of Muscle Coordination | • Going to the bathroom<br>• Driving |
| **Speech Disturbance** | Speech is assessed during normal conversation. | Assessment of neurological function that speech | • Dysarthria<br>• Dysmetria<br>• Dysphasia<br>• Ataxia<br>• Dystonia<br>• Dyskinesias<br>• Hypotonia<br>• Lower facial weakness/ muscle weakness<br>• Slurred speech<br>• Loss of Muscle Coordination | • Socializing<br>• Working<br>• Having a conversation<br>• Talking on the phone<br>• Communicating with caregiver or family |
| **Finger chase test** | Patient sits comfortably. If necessary, support of feet and trunk is allowed.<br>Examiner sits in front of patient and performs 5 consecutive sudden | Assessment of neurological function that measures smooth, coordinated upper-extremity movement, tremor, and accuracy of fine motor function/target | • Dysmetria<br>• Tremor<br>• Dyskinesias<br>• Ataxia<br>• Spasticity | • Shaving<br>• Using computer mouse /keyboard<br>• Using smartphone/ Dialing phone/Texting<br>• Going to the bathroom (i.e. buttoning pants)<br>• Dressing |
|  | and fast pointing movements in unpredictable directions in a front plane, at about 50% of patient's reach. Movements have an amplitude of 30 cm and a frequency of 1 movement every 2 s. Patient is asked to follow the movements with index finger, as fast and precisely as possible. | accuracy. | • Dystonia<br>• Hypotonia<br>• Muscle weakness<br>• Loss of Muscle Coordination | • Driving<br>• Food preparation (i.e. cannot lift silverware or a glass to mouth accurately/ feed oneself / serve self food)<br>• Writing/ writing legibly<br>• Turning lock in key<br>• Self care (i.e. Cannot brush teeth, cannot put on makeup)<br>• Sewing/needle craft/ handling tools<br>• Car repairs; home repairs<br>• Play instrument |
| **Nose-finger test** | Patient sits comfortably. If necessary, support of feet and trunk is allowed.<br>Patient is asked to point repeatedly with index finger from their nose to the examiner's finger, which is in front of the patient at about 90% of the patient's reach. Movements are performed at moderate speed. | Assessment of neurological function that measures smooth, coordinated upper-extremity movement, tremor, and accuracy of fine motor function/target accuracy. | • Dysmetria<br>• Tremor<br>• Dyskinesias<br>• Hypotonia<br>• Dystonia<br>• Muscle weakness<br>• Loss of Muscle Coordination<br>• Ataxia<br>• Spasticity |  |
| **Fast alternating hand movement** | Patient sits comfortably. If necessary, support of feet and trunk is allowed.<br>Patient is asked to perform 10 cycles of repetitive alternation of pro- and supinations of the hand on their thigh as fast and precise as possible. | Assessment of neurological function that *measures* several aspects of coordination; when a patient has neurological dysfunction, one movement often cannot be quickly followed by its opposite (e.g. movement is not synchronous) and movements are slow, irregular, and clumsy. | • Dysdiadochokinesia<br>• Slowing of rapid alternating movements (e.g. due to pyramidal dysfunction)<br>• Ataxia<br>• Dysmetria<br>• Muscle weakness<br>• Loss of Muscle Coordination<br>• Hypotonia<br>• Dyskinesias<br>• Dystonia<br>• Tremor<br>• Spasticity | • Cannot hand things to another person<br>• Unable to turn pages<br>• Cannot open door/ turn doorknob<br>• Inability to lift objects<br>• Brushing Teeth<br>• Turn key in lock/ a door nob<br>• Knife skills/ preparing food<br>• Sports<br>• Handshakes |
| **Heel-shin slide** | Patient lies on examination bed, without sight of their legs. Patient is asked to lift one leg, point with the heel to the opposite knee, slide down along the shin to the angle, and lay the leg back on the examination bed. The task is performed 3 times. Slide-down movements should be performed within 1 s. | Assessment of lower limb coordination that measures, smooth, coordinated, precise lower-extremity movement. | • Dysmetria<br>• Dyskinesias<br>• Ataxia<br>• Dystonia<br>• Muscle weakness<br>• Loss of Muscle Coordination<br>• Hypotonia<br>• Tremor<br>• Spasticity | • Cannot drive safely<br>• Cannot take shoes on and off<br>• Cannot stand alone in shower<br>• Impairs ability to walk safely/ affects balance |

### SARA for Non-Ataxia Disorders

The SARA scale was thus initially developed to measure symptoms of cerebellar ataxia in dominant spinocerebellar ataxia (SCA). Later, it was validated for use in other various types of ataxias [4, 4, 10]. More recently, the SARA has been increasingly utilized as a clinical outcome assessment for a wide range of disorders, ranging from rare entities such as lysosomal disorders to more common pediatric cancers [11–14]. The generalizability of the SARA may be related to multi-item assessments that can be categorized into 4 disease-agnostic functionally different categories:

A. Ambulation & function of lower extremities: Test items (1) Gait, (8) heel-shin slide
B. Postural balance: Test items (2) Stance, (3) Sitting
C. Speech: Test items (4) Speech disturbance
D. Function of upper extremities (Fine motor): Test items (5) Finger chase, (6) Nose-finger, (7) Fast alternating hand movements

When a patient performs voluntary movements as part of the SARA assessments, such as speaking or walking, this requires a sequence of coordinated actions (e.g., adequate motivation, attention, cognition, hearing, planning of movements, muscle power, strength, control and precision of movements) that involve many regions of the brain from the frontal cortex, somatosensory cortex, basal ganglia, cerebellum, brainstem to the corticospinal tract, and the spinal cord. In LSDs, cellular damage and cell death occur throughout the entirety of the central nervous system, manifesting as a wide range of heterogeneous neurological signs and symptoms (e.g., dysarthrophonia, ocular motor, dysmetria, ataxia, dysdiadochokinesia, dystonia, tremor, hypotonia, dyskinesias, spasticity – see **Table 2**), each of which could impact the ability of the patient to undertake the necessary sequence and precision of actions required to perform the SARA tasks, ultimately resulting in dysfunction in one or more of the above functional categories.

**Table 2:** Neurological Symptoms of Lysosomal Storage Disorders/ Impact on SARA Test Item.

| Region of Brain | Specific Neurological Symptom | Description (if applicable) | SARA Item Directly Affected by the Specific Symptom |
| --- | --- | --- | --- |
| **Cerebellum [22]** | Ataxia | Lack of precision in voluntary muscle movements and coordination and sequence of movements | All items (gait, speech disturbance, finger chase, nose-finger test, fast alternating hand movement, and heel-shin slide) |
|  | Dysarthria | Motor speech disorder due to impairments in the muscular control of speech which can affect the strength, speed, range, tone, and accuracy of the speech | Speech |
|  | Dysmetria | Inability to control the distance, speed, and range of motion necessary to perform smoothly-coordinated movements | All items (gait, speech disturbance, finger chase, nose-finger test, fast alternating hand movement, and heel-shin slide)<br>*Note: diagnostic tests for Dysmetria specifically include the nose-finger test and heel-shin slide)* |
|  | Dysphagia | Difficulty swallowing (which also makes it difficult to eat, drink, and speak) | Speech |
|  | Dysdiadochokinesia | Inability to perform rapid alternating muscle movements (opening and closing fists, tap shoe, alter hands) | Fast alternating hand movement<br>*Note: the diagnostic test for Dysdiadochokinesia is the fast alternating hand movement test)* |
| **Basal Ganglia [23]** | Dystonia | Involuntary muscle co-contractions | All items (gait, speech disturbance, finger chase, nose-finger test, fast alternating hand movement, and heel-shin slide) |
|  | Tremor | Involuntary quivery movement | Finger chase, nose-finger test, fast alternating hand movement (all items may be impacted if essential tremor) |
|  | Dyskinesias | Involuntary, erratic, writhing movements (tics, tremors, shakes, full body movements) | All items (gait, speech disturbance, finger chase, nose-finger test, fast alternating hand movement, and heel-shin slide) |
|  | Hypotonia | Low (decreased) muscle tone | All items (gait, speech disturbance, finger chase, nose-finger test, fast alternating hand movement, and heel-shin slide) |
| **Pyramidal tract [24]** | Spasticity | Muscle control disorder that causes abnormal muscle tightness, stiffness, or pull | Gait, sitting, stance, finger chase, nose-finger test, fast alternating hand movement, and heel-shin slide |
|  | Slowing of rapid alternating movements | N/A | Fast alternating hand movement and heel-shin slide |
|  | Lower facial weakness and changes to speech | N/A | Speech |
| **Brain Stem [25]** | Balance Problems | Muscle control disorder that causes abnormal muscle tightness, stiffness, or pull | Gait, sitting, and stance |
|  | Muscle Weakness | N/A | All items (gait, speech disturbance, finger chase, nose-finger test, fast alternating hand movement, and heel-shin slide) |
|  | Slurred Speech | N/A | Speech |
|  | Loss of Muscle Coordination | N/A | All items (gait, speech disturbance, finger chase, nose-finger test, fast alternating hand movement, and heel-shin slide) |

Therefore, we hypothesized that a change in the functional performance as assessed by the SARA scale could be indicative of broad alterations in many functional neurological networks, allowing its use as a measure of overall neurological disease severity in LSDs, as opposed to an isolated measure of cerebellar ataxia.

## Methods

### Study Objective

Given the increased use of the SARA scale as an endpoint for LSDs, we aimed to evaluate the reliability, reproducibility, and responsiveness of the scale for LSDs that feature central nervous system involvement and investigate the range of neurological signs and symptoms which could be captured and measured. The study also evaluated a minimum threshold of change which would demonstrate clinical and functional significance.

### Participants

Data was analyzed from three clinical trials conducted with the agent N-acetyl-L-leucine (IB1001) for LSDs, including 2 Phase IIb, open-label, rater-blinded studies with Niemann-Pick disease type C (NPC) [“IB1001-201”, NCT03759639, n= 32 patients] and GM2 Gangliosidoses (Tay Sachs and Sandhoff diseases) [“IB1001-202”, NCT03759665, n=30 patients] and a Phase III, double-blind, placebo-controlled trial for NPC [“IB1001-301”, NCT05163288, n=60]. In the IB1001-201 and IB1001-202 studies, the SARA was a secondary endpoint; in the IB1001-301 study, the SARA was the primary endpoint.

Patients were recruited in the three clinical trials between 07-Jun-2019 and 22-Dec-2022 from 17 centers.

This study was conducted in accordance with the International Conference for Harmonisation (of Technical Requirements for Pharmaceuticals for Human Use) - Good Clinical Practice Guideline, the General Data Protection Regulator, and the Declaration of Helsinki. Approval was obtained by the applicable responsible central research ethics committees / institutional review boards for each center. Written informed consent was obtained from all study participants (or their parent/ legal representative) at enrolment. The methodology and results of each trial has been previously published [11–13, 15, 16].

### Procedures

The study design/schema for the Phase IIb (IB1001-201, IB1001-202) and Phase III (IB1001-301) trials are presented in **Figure 1A & B** [11–13, 15, 16].

**Fig. 1.**
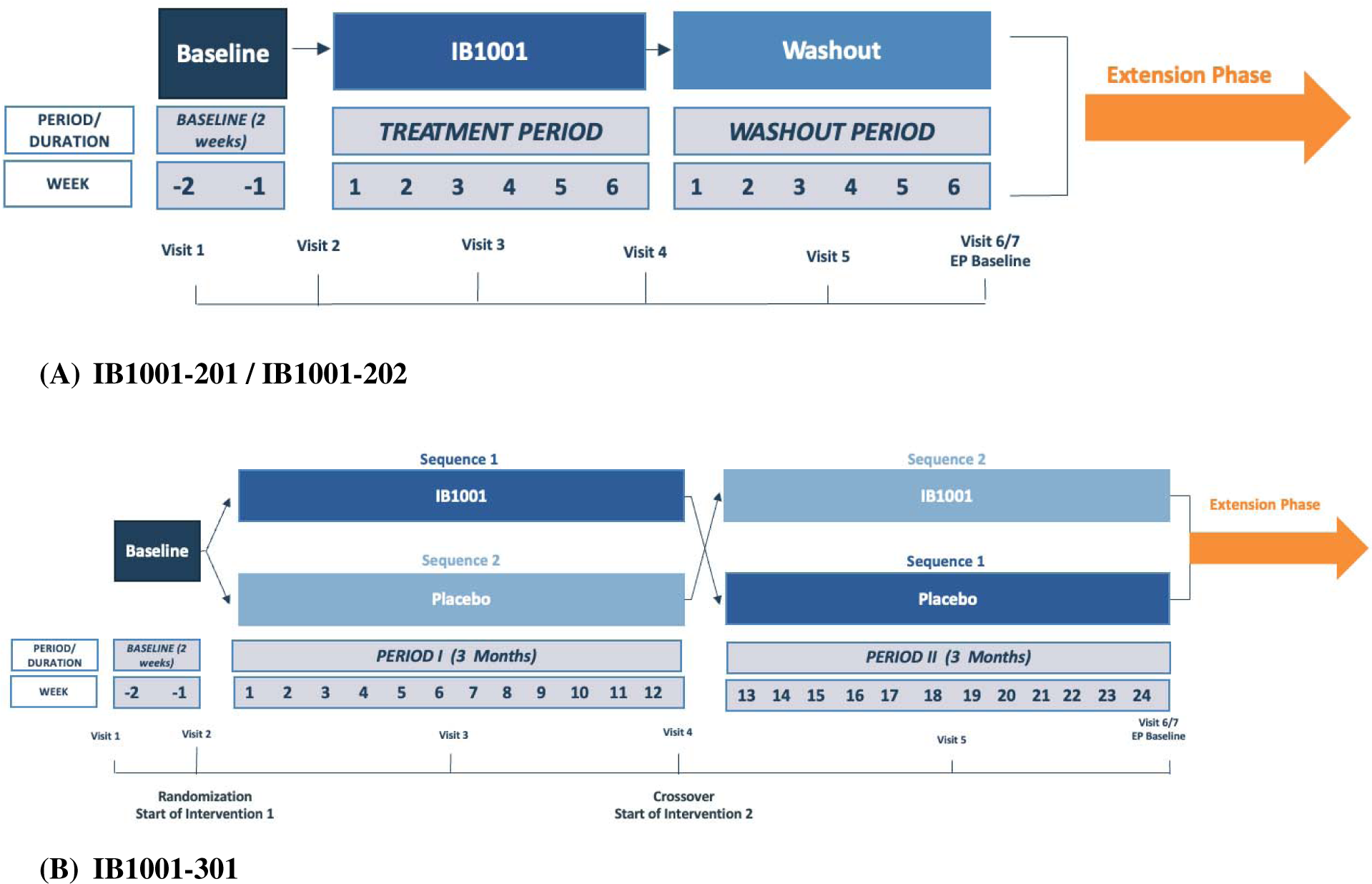
Study Schemes from IB1001 Clinical Trials.

In each study, the SARA was assessed by a qualified investigator at every study visit. The investigators underwent standardized training on the assessment and the same investigator was required to perform the SARA assessment at each visit for each patient to exclude confounding by inter-rater variability. For the IB1001-201 and IB1001-202 studies, this included two baseline visits at approximately Day 1 (Visit 1) and after 2 weeks of screening (Visit 2), two treatment visits conducted after approximately 4 weeks (Visit 3) and 6 weeks (Visit 4) of treatment with IB1001, and 2 washout visits conducted after approximately 4 weeks (Visit 5) and 6 weeks (Visit 6) of post-treatment from IB1001. In the IB1001-301 study, this included two baseline visits at approximately Day 1 (Visit 1) and after 2 weeks of screening (Visit 2), two treatment visits conducted after approximately 6 weeks (Visit 3) and 12 weeks (Visit 4) of treatment with IB1001 or Placebo, and two treatment visits conducted after approximately 6 weeks (Visit 5) and 12 weeks (Visit 6) of the opposite treatment (IB1001 or Placebo).

In addition, the Clinical Global Impression of Improvement was assessed by the Investigator, Caregiver, and Patient (if able) at the end of every treatment period, e.g. at Visit 4 (versus Visit 2) and Visit 6 (versus Visit 4) [17]. Finally, in the Phase III trial, exit interviews (in the form of semi-structured interviews) were conducted with the patient (if able) and/or caregiver (if applicable) at Visit 6 (end of the Parent Study) or the ET visit to better inform and evaluate the meaningfulness of the within-patient changes on the outcome assessments and document the relevance and meaningfulness of functional improvements in patients’ everyday lives (see the questionnaire in **Supplemental Material 1**) [18]. These exit interviews were conducted prior to any unblinding.

### Statistical Analyses

#### Reliability & Reproducibility

In each clinical trial, participants were assessed twice during the baseline period (before any intervention) at visits approximately 14 – 21 days apart (Visit 1 and Visit 2). The mean and (SD) were computed for each of the baseline visits as well as the difference between Visits 1 and Visit 2 for each trial. The results for the three trials were compared for consistency and two-sided t-tests were used to test for group differences between the 301 trial and the 201 and 202 trials. In addition, the mean (SD) and the difference between the baseline Visit 1 and Visit 2 for patients <10 years old were computed to assess the reliability of the SARA assessment in these younger patients. Given the small sample sizes in the three trials, the data was combined to enable statistical interpretation. These results were compared to the results of patients aged 10 years and older for consistency and a two-sided t-test was used to test for group differences.

To determine the test re-test reliability of SARA, inter-class correlations (ICCs) were calculated from all three trials for the total SARA score between baseline Visit 1 and Visit 2, and also by each of the 8 items (e.g., SARA gait Visit 1 versus Visit 2). For the IB1001-201 and IB1001-202 trials, the mean and standard deviation (SD) were also computed for the end of post-treatment washout Visit (6), and the mean (SD) difference between Visit 1 and Visit 6 was calculated. The results were compared for consistency and an independent sample t-test was used to test for group differences between the two studies. A calculation of the mean (SD) difference was also calculated for the subgroup of patients aged <10 years from the IB1001-201 and IB1001-202 trials. These results were compared to the results of patients aged 10 years and older for consistency and a two-sided t-test was used to test for group differences. The ICCs between the Visit 1 (baseline) and Visit 6 (post-treatment washout) scores were also computed for the total SARA scale as well as for the 8 individual test items (gait, stance, sitting, speech disturbances, finger-chase, nose-finger, fast alternating hand movement, heel-shin slide).

Coefficients exceeding 0.80 were considered acceptable for the total SARA scale; coefficients above 0.70 were considered acceptable for each of the 8 single SARA items [19].

#### Responsiveness

The responsiveness of the SARA scale was defined as the ability to detect clinically important changes. To assess this, for the IB1001-301 trial data, SARA scores were compared with Clinical Global Impression of Improvement (CGI-I) scores in order to determine whether changes in SARA reflect the clinical changes recorded by Investigators, Caregivers and Patients.

This analysis was carried out for the second treatment period of the IB1001-301 trial (the IB1001-201 and IB1001-202 trials were open-label and therefore not considered appropriate for comparison; in the IB1001-301 trial treatment period 1, 50% of patients were on placebo treatment and therefore this period was not appropriate for comparison). Responsiveness was defined as the ability to categorize patients as Improved (patients rated as minimally, much, or very much improved) or Unchanged (no change) or Worsened (minimally, much, or very much worse) as a function of Δ SARA with re-scored CGI-I as external criteria. Visit 4 (end of treatment with IB1001 or Placebo) to Visit 6 (end of opposite treatment) changes in SARA (Δ SARA) scores were compared with CGI-I at Visit 6. Approximately half the patients – those randomized in the sequence IB1001-Placebo – were expected to show stable or worsening SARA scores, and the other sequence – Placebo-IB1001 – to register improvements.

The CGI-I scale has been shown to be able to successfully differentiate between responders and non-responders to investigational study drugs [20]. Accordingly, CGI-I scores were allocated to categories “Improved”, “Unchanged” and “Worsened”. The mean and 95% CI for the no-change group were calculated. The mean values of ΔSARA for the improved and worsened categories were calculated and compared with the no change 95% CI to determine whether the mean change of either group overlapped with the 95% CI for the no-change group.

To further quantify the ability of SARA to assess clinically meaningful improvements, a second analysis was carried out on the CGI-I data. Here, CGI-I data was further collapsed into the binary categories “Improved” (minimally, much, or very much improved) and “Not Improved” (no change, minimally, much, or very much worse) For each CGI-I, the Confusion Matrix was computed at each value of Δ SARA, and a Receiver Operating Characteristic (ROC) curve was calculated for the True Positive Rate as a function of the False Positive Rate [FDA Guidance for Industry 2023]. The area under the curve (AUC), equating to the ability to detect a clinical change, was calculated. An AUC value greater than 0.70 was considered the minimum threshold for discriminative ability [19, 21].

#### Correlation Between Total SARA Score and Activities of Daily Life

To assess whether changes on the SARA scale correlated with patient/caregiver-reported clinically meaningful improvements in everyday function, activities of daily life, and/or quality of life, the exit interviews from the IB1001-301 study were qualitatively assessed. For patients who experienced a 1-point or greater improvement on the SARA scale after treatment with IB1001 or, for those randomized to receive IB1001 followed by Placebo, a 1-point worsening or greater on the SARA, a qualitative system review aggregated their exit interviews into “responders” versus “non-responders”. “Responders” were defined as exit interviews where the patient/caregiver described the changes during the IB1001-301 clinical trial to be beneficial, and reported improvement in everyday function and quality of life which were considered to be clinically meaningful. Content Analysis was applied to determine if the reported changes were limited to the symptom of cerebellar ataxia, or if changes in other neurological signs and symptoms could potentially be associated with changes on the SARA scale.

## Results

### Patients

The analyzed subset of 122 patients (mean age 27.1, range 5 – 67, years; 66 male, 56 female, 81 NPC, 30 GM2) had a mean SARA score of 15.17 (7.28) at Visit 1. As indicated by the distribution of the SARA baseline scores (SD 7.28, range 4.5 to 35), the cohort was representative of a broad range of disease severity except asymptomatic patients or the most severely impaired patients. In total, 122 Δ SARA scores from Visit 1 and Visit 2 (from the IB1001-201, IB1001-202, and IB1001-301 study) and 57 Δ SARA scores from Visit 1 and Visit 6 (from the IB1001-201 and IB1001-202 study) were evaluated for reliability and reproducibility. 58 Investigator CGI-I scores, 50 Caregiver CGI-I scores, and 49 Patient CGI-I scores comparing Visit 4 to Visit 6 (from the IB1001-301 study) and were evaluated against the corresponding 58 Δ SARA scores for responsiveness. Across the three trials, there were 15 patients aged <10 years (range 5 to 9 years); in the IB1001-201 and IB1001-202 trials there were 7 patients aged <10 years (range 6 to 9 years).

### Reliability & Reproducibility

Test re-test data for all three trials for the baseline period (Visit 1 and Visit 2) are shown in **Table 3**. The mean (SD) change between baseline Visit 1 and Visit 2 for the IB1001-201, IB1001-202 and IB1001-301 trials were -0.30 (1.75), +0.02 (1.23), and +0.03 (1.96) respectively (Table 2). There was no statistically significant change in the mean value from Visit 1 to Visit 2 for any study or statistically meaningful difference found between the re-test IB1001-301 score and the IB1001-201 or IB1001-202 scores, reflecting the SARA was highly reliable/reproducible. For patients <10 years, the mean (SD) change from baseline Visit 1 and Visit 2 was 0.27 (2.05) which was not statistically significantly different from the cohort of patients aged 10 years and older (p=0.50), reflecting the SARA was also reliable/reproducible in this population.

**Table 3.**
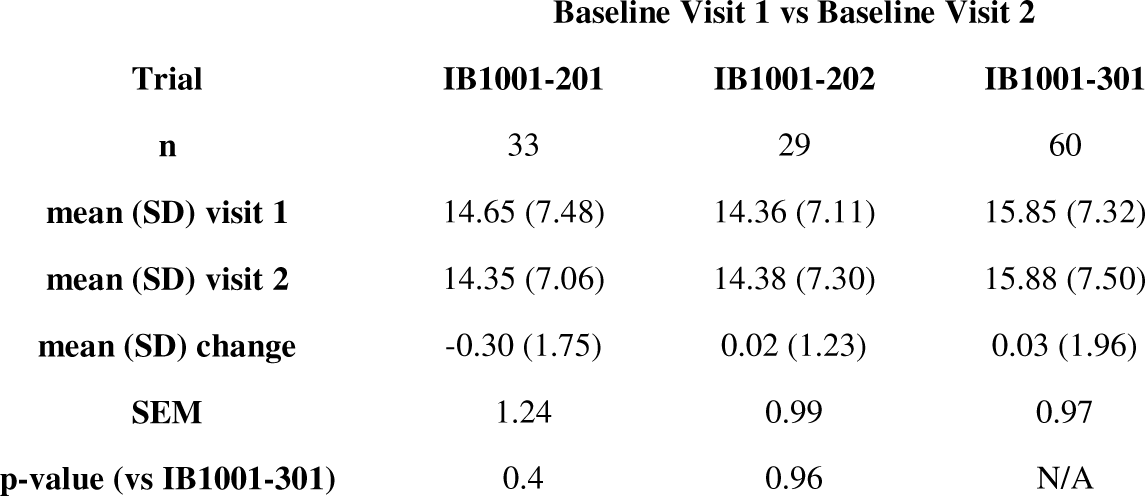
Test re-test data Baseline Visit 1 & Baseline Visit 2.

The ICCs for the SARA scale and each of its 8 items are given in **Table 4**. The SARA scale correlations were 0.971, 0.986 and 0.966 for the three trials. These item-level correlations all exceeded the 0.70 threshold and showed strong agreement, demonstrating a high degree of internal consistency. The Sitting test ICC was 1.0 for the IB1001-202 which was an indication of the flooring effect for that item (20 of 29 scores were 0).

**Table 4.** Interclass Correlations between Baseline Visit 1 & Baseline Visit 2 (approximately 2-3 weeks apart)

|  | Trial |  |  |
| --- | --- | --- | --- |
|  | IB1001-201 | IB1001-202 | IB1001-301 |
| *Total Score* |  |  |  |
| SARA | 0.971 | 0.986 | 0.966 |
| *Individual Test Item* |  |  |  |
| Gait | 0.973 | 0.988 | 0.965 |
| Stance | 0.943 | 0.945 | 0.895 |
| Sitting | 0.919 | 1 | 0.898 |
| Speech | 0.923 | 0.94 | 0.933 |
| Nose Finger | 0.756 | 0.963 | 0.882 |
| Finger Chase | 0.857 | 0.902 | 0.793 |
| Hand Movements | 0.94 | 0.9 | 0.887 |
| Heel Shin | 0.976 | 0.927 | 0.921 |

Test re-test data for the IB1001-201 and IB1001-202 trials for the baseline versus post-treatment washout period data (Visit 1 and Visit 6) are shown in **Table 5**. There were no differences observed between the baseline visit and the post-washout visit (Visit 1 to Visit 6); the mean change (SD) in SARA score was -0.03 (2.61) in the IB1001-201 study and -0.04 (2.07) in the IB1001-202 study. This further reinforced the reliability of the administration of the SARA scale, and also demonstrated the absence of a learning effect on the 8 SARA items. For patients <10 years, the mean (SD) change from baseline Visit 1 and post-treatment washout Visit 6 was 0.00 (1.61) which was not statistically significantly different from the cohort of patients aged 10 years and older (p=0.95), reflecting the SARA was also reliable/reproducible in this population.

**Table 5.** Test re-test data Baseline Visit 1 & Post-Treatment Washout Visit 6 (approximately 14 weeks apart)

|  | Trial |  |
| --- | --- | --- |
|  | IB1001-201 | IB1001-202 |
| n | 31 | 26 |
| mean (SD) visit 1 | 14.94 (7.64) | 14.19 (7.48) |
| mean (SD) visit 6 | 14.90 (8.16) | 14.15 (8.31) |
| mean (SD) change | -0.03 (2.61) | -0.04 (2.07) |
| SEM | 0.97 | 0.97 |
| p-value | 0.99 |  |

The ICCs for this comparison are shown in **Table 6**. The total SARA scale ICCs were high and above the 0.80 threshold. These item-level correlations all exceeded the 0.70 thresholds and showed strong agreement. There was also further evidence of the flooring effect in the Sitting test where the IB1001-202 trial ICC was 1.0.

**Table 6.** Interclass Correlations between Baseline Visit 1 & Post-Treatment Visit 6.

|  | Trial |  |
| --- | --- | --- |
|  | IB1001-201 | IB1001-202 |
| *Total Score* |  |  |
| **SARA** | 0.947 | 0.967 |
| *Individual Test Item* |  |  |
| **Gait** | 0.942 | 0.894 |
| **Stance** | 0.888 | 0.854 |
| **Sitting** | 0.839 | 1 |
| **Speech** | 0.731 | 0.896 |
| **Nose Finger** | 0.826 | 0.889 |
| **Finger Chase** | 0.62 | 0.767 |
| **Hand Movements** | 0.781 | 0.894 |
| **Heel Shin** | 0.878 | 0.972 |

### Responsiveness

The CGI-I scores categorized as “unchanged”, “improved” and “worsened” and corresponding Δ SARA values for the IB1001-301 trial (Visit 4 to Visit 6) are summarized in **Table 7**. Δ SARA values ranged between -5.5 and +6.5 (expected variance given approximately 50% of patients in this treatment period were commencing IB1001, and 50% were stopping IB1001 treatment). The Mean (SD) was -0.06 (2.72).

**Table 7.** Patient count and mean ΔSARA for the collapsed CGI-I assessment categories. The results were studied to determine whether the mean change of either the Improved and Worsened group overlapped with the 95% CI for the Unchanged group for each of the three assessor groups: Investigator, Caregiver and Patient.

| CGI-I | CGI-I improved |  | CGI-I unchanged |  | CGI-I worsened |  |
| --- | --- | --- | --- | --- | --- | --- |
| | n | mean ΔSARA (95% CI) | n | mean ΔSARA (95% CI) | n | mean ΔSARA (95% CI) |
| **Physician** | 24 | -1.79 (-2.71, -0.88) | 16 | -0.09 (-0.83, 0.64) | 18 | 1.16 (0.05, 2.27) |
| **Caregiver** | 20 | -1.18 (-2.53, 0.18) | 14 | -0.07 (-1.30, 1.15) | 16 | 0.78 (-0.50, 2.07) |
| **Patient** | 15 | -1.90 (-3.20, -0.60) | 22 | 0.41 (-0.81, 1.62) | 12 | 0.87 (-0.65, 2.38) |

The CGI-I values categorized as “improved” or “not improved” used for the AUC calculations are summarized in Table 8. As described above, patients randomized in the sequence Placebo-IB1001 were expected to show improvement during this period those randomized in the sequence IB1001-Placebo were expected to show worsening during the second period if the patient was a responder to the study drug. The AUC for Investigator CGI-I was 0.82, for Caregiver CGI-I it was 0.71, and for Patient CGI-I AUC was 0.77. All CGI-Is were above the threshold for discriminative ability, supporting changes in SARA aligned with CGI-I assessments of changes in patients’ overall function and well-being.

**Table 8.** Patient count for CGI-I collapsed to a binary classifier “Improved” or “Not Improved” for each of the three assessor groups: Investigator, Caregiver and Patient.

|  | CGI-I<br>improved | CGI-I Not<br>Improved |
| --- | --- | --- |
|  | n | n |
| Investigator | 24 | 34 |
| Caregiver | 20 | 30 |
| Patient | 15 | 34 |

### Correlation Between Total Score and Activities of Daily Life

42 exit interviews were qualitatively assessed for patients who experienced a 1-point or greater improvement on the SARA scale after treatment with IB1001 or, for those randomized to receive IB1001 followed by Placebo, a 1-point worsening or greater on the SARA. 70% of patients were identified to be responders to the study drug, meaning that the patient/caregiver described clinically meaningful, relevant changes in exit interviews, reinforcing previous findings that there is a close correlation between total SARA score and measures of daily activities and functional assessment and that a minimum 1-point change is clinically meaningful [2]. The exit interviews further elucidated that clinically meaningful changes included: increase in strength and energy; improved cataplexy, dysphagia, ataxia, dystonia; reduced (less) pain in muscles/general; improved speech, more easily understood/fluent speech, easier to integrate into a conversation, easier to communicate with; improved ambulation, mobility, balance, coordination, and autonomous gait; reduced falls; improved fine motor skills/general motor skills, less tremor; improved cognition, concentration, brain fog, focus, memory, cooperation, behavior, mood; reduced anxiety; less swallowing problems, less coughing while swallowing; improved incontinence (urine and anal); reduced seizures; improved sleep; improved ability to perform everyday tasks (feeding, dressing, playing, work, following orders, participating in leisure activities), and were not limited to the isolated measure of cerebellar ataxia. Examples from the exit interviews are provided in **Table 9**.

**Table 9.**
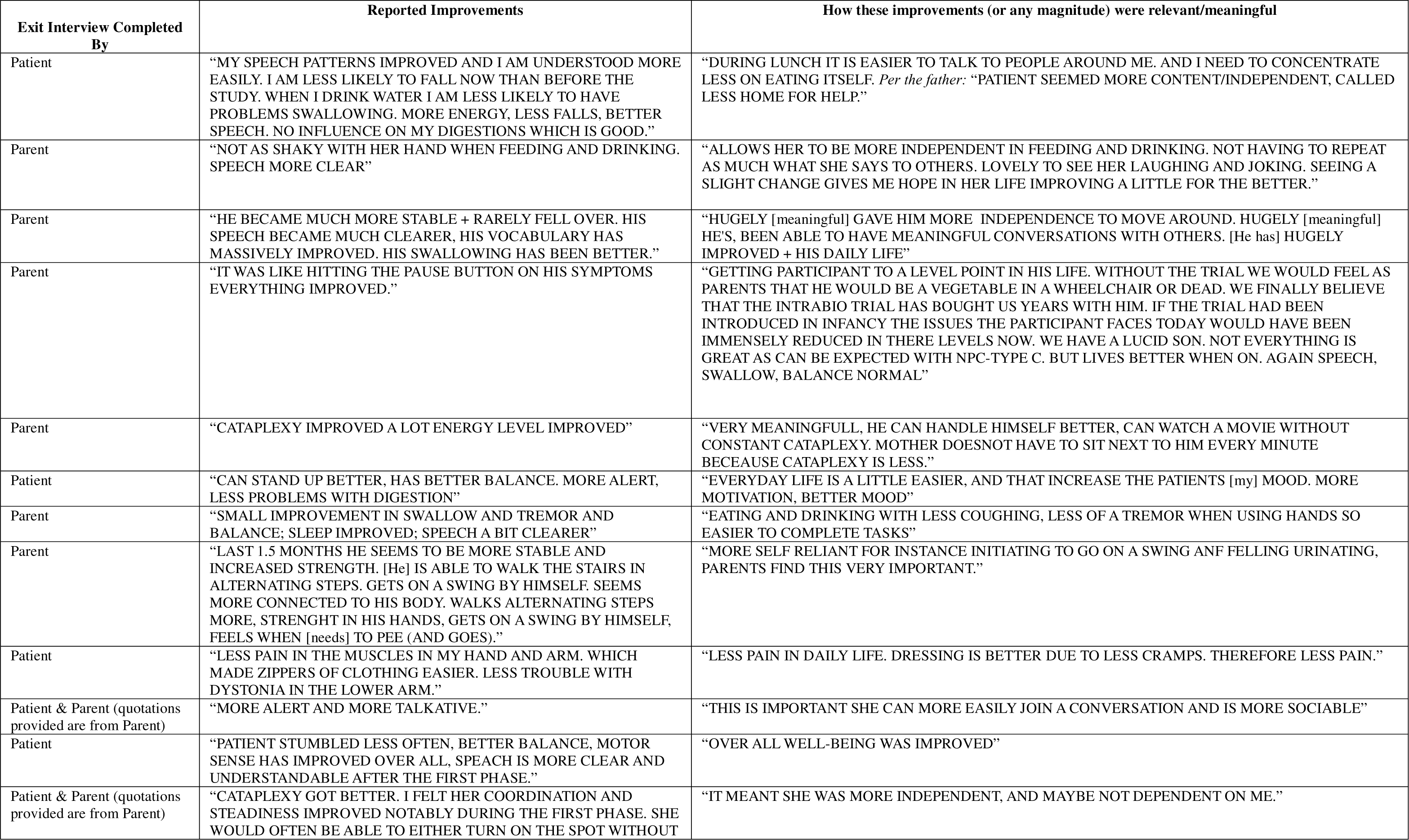
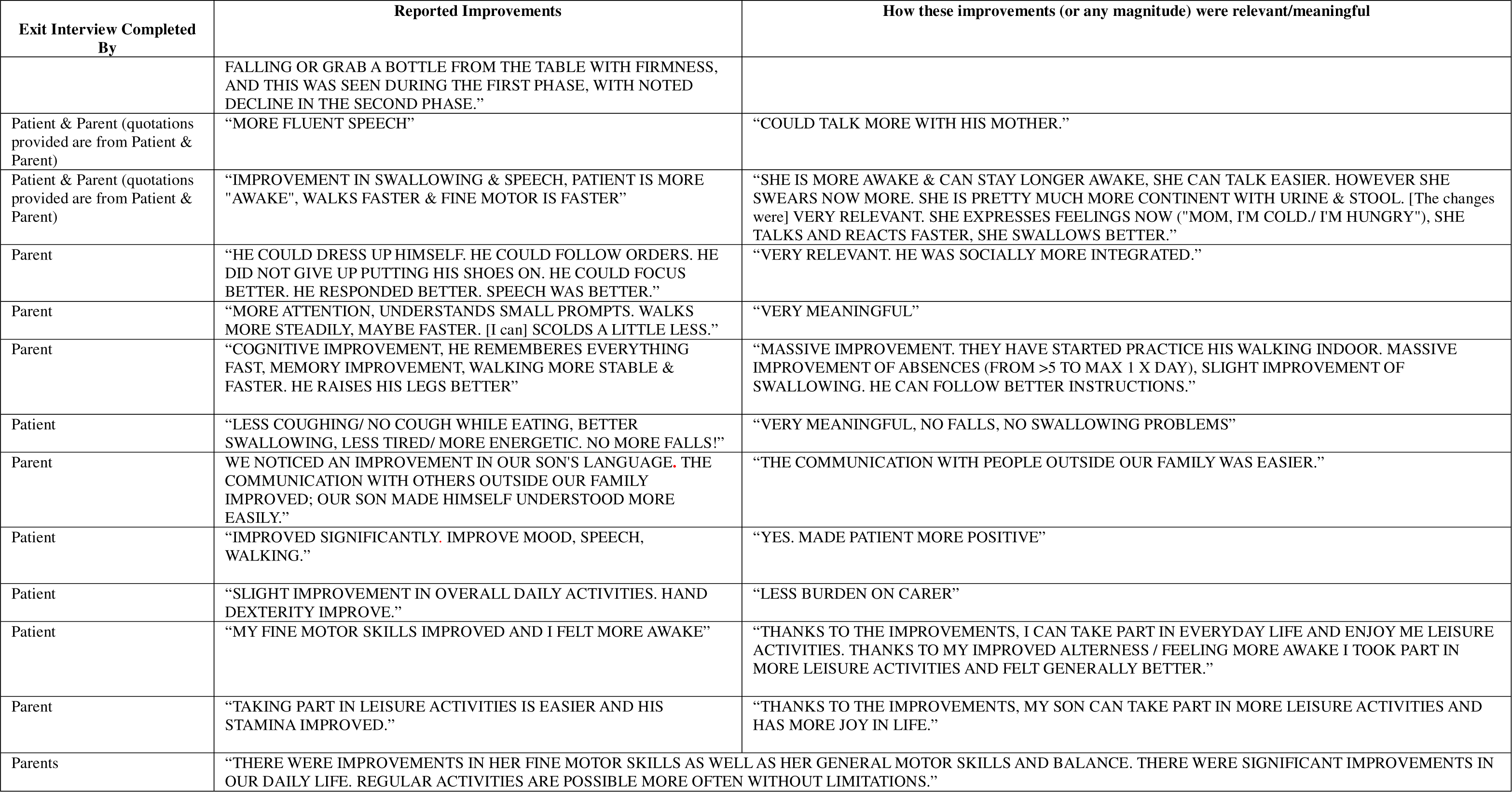
Exit Interview Responses.

| Exit Interview Completed By | Reported Improvements | How these improvements (or any magnitude) were relevant/meaningful |
| --- | --- | --- |
| Patient | “MY SPEECH PATTERNS IMPROVED AND I AM UNDERSTOOD MORE EASILY. I AM LESS LIKELY TO FALL NOW THAN BEFORE THE STUDY. WHEN I DRINK WATER I AM LESS LIKELY TO HAVE PROBLEMS SWALLOWING. MORE ENERGY, LESS FALLS, BETTER SPEECH. NO INFLUENCE ON MY DIGESTIONS WHICH IS GOOD.” | “DURING LUNCH IT IS EASIER TO TALK TO PEOPLE AROUND ME. AND I NEED TO CONCENTRATE LESS ON EATING ITSELF. *Per the father:* “PATIENT SEEMED MORE CONTENT/INDEPENDENT, CALLED LESS HOME FOR HELP.” |
| Parent | “NOT AS SHAKY WITH HER HAND WHEN FEEDING AND DRINKING. SPEECH MORE CLEAR” | “ALLOWS HER TO BE MORE INDEPENDENT IN FEEDING AND DRINKING. NOT HAVING TO REPEAT AS MUCH WHAT SHE SAYS TO OTHERS. LOVELY TO SEE HER LAUGHING AND JOKING. SEEING A SLIGHT CHANGE GIVES ME HOPE IN HER LIFE IMPROVING A LITTLE FOR THE BETTER.” |
| Parent | “HE BECAME MUCH MORE STABLE + RARELY FELL OVER. HIS SPEECH BECAME MUCH CLEARER, HIS VOCABULARY HAS MASSIVELY IMPROVED. HIS SWALLOWING HAS BEEN BETTER.” | “HUGELY [meaningful] GAVE HIM MORE INDEPENDENCE TO MOVE AROUND. HUGELY [meaningful] HE'S, BEEN ABLE TO HAVE MEANINGFUL CONVERSATIONS WITH OTHERS. [He has] HUGELY IMPROVED + HIS DAILY LIFE” |
| Parent | “IT WAS LIKE HITTING THE PAUSE BUTTON ON HIS SYMPTOMS EVERYTHING IMPROVED.” | “GETTING PARTICIPANT TO A LEVEL POINT IN HIS LIFE. WITHOUT THE TRIAL WE WOULD FEEL AS PARENTS THAT HE WOULD BE A VEGETABLE IN A WHEELCHAIR OR DEAD. WE FINALLY BELIEVE THAT THE INTRABIO TRIAL HAS BOUGHT US YEARS WITH HIM. IF THE TRIAL HAD BEEN INTRODUCED IN INFANCY THE ISSUES THE PARTICIPANT FACES TODAY WOULD HAVE BEEN IMMENSELY REDUCED IN THERE LEVELS NOW. WE HAVE A LUCID SON. NOT EVERYTHING IS GREAT AS CAN BE EXPECTED WITH NPC-TYPE C. BUT LIVES BETTER WHEN ON. AGAIN SPEECH, SWALLOW, BALANCE NORMAL” |
| Parent | “CATAPLEXY IMPROVED A LOT ENERGY LEVEL IMPROVED” | “VERY MEANINGFULL, HE CAN HANDLE HIMSELF BETTER, CAN WATCH A MOVIE WITHOUT CONSTANT CATAPLEXY. MOTHER DOESNOT HAVE TO SIT NEXT TO HIM EVERY MINUTE BECEAUSE CATAPLEXY IS LESS.” |
| Patient | “CAN STAND UP BETTER, HAS BETTER BALANCE. MORE ALERT, LESS PROBLEMS WITH DIGESTION” | “EVERYDAY LIFE IS A LITTLE EASIER, AND THAT INCREASE THE PATIENTS [my] MOOD. MORE MOTIVATION, BETTER MOOD” |
| Parent | “SMALL IMPROVEMENT IN SWALLOW AND TREMOR AND BALANCE; SLEEP IMPROVED; SPEECH A BIT CLEARER” | “EATING AND DRINKING WITH LESS COUGHING, LESS OF A TREMOR WHEN USING HANDS SO EASIER TO COMPLETE TASKS” |
| Parent | “LAST 1.5 MONTHS HE SEEMS TO BE MORE STABLE AND INCREASED STRENGTH. [He] IS ABLE TO WALK THE STAIRS IN ALTERNATING STEPS. GETS ON A SWING BY HIMSELF. SEEMS MORE CONNECTED TO HIS BODY. WALKS ALTERNATING STEPS MORE, STRENGHT IN HIS HANDS, GETS ON A SWING BY HIMSELF, FEELS WHEN [needs] TO PEE (AND GOES).” | “MORE SELF RELIANT FOR INSTANCE INITIATING TO GO ON A SWING ANF FELLING URINATING, PARENTS FIND THIS VERY IMPORTANT.” |
| Patient | “LESS PAIN IN THE MUSCLES IN MY HAND AND ARM. WHICH MADE ZIPPERS OF CLOTHING EASIER. LESS TROUBLE WITH DYSTONIA IN THE LOWER ARM.” | “LESS PAIN IN DAILY LIFE. DRESSING IS BETTER DUE TO LESS CRAMPS. THEREFORE LESS PAIN.” |
| Patient & Parent (quotations provided are from Parent) | “MORE ALERT AND MORE TALKATIVE.” | “THIS IS IMPORTANT SHE CAN MORE EASILY JOIN A CONVERSATION AND IS MORE SOCIABLE” |
| Patient | “PATIENT STUMBLED LESS OFTEN, BETTER BALANCE, MOTOR SENSE HAS IMPROVED OVER ALL, SPEACH IS MORE CLEAR AND UNDERSTANDABLE AFTER THE FIRST PHASE.” | “OVER ALL WELL-BEING WAS IMPROVED” |
| Patient & Parent (quotations provided are from Parent) | “CATAPLEXY GOT BETTER. I FELT HER COORDINATION AND STEADINESS IMPROVED NOTABLY DURING THE FIRST PHASE. SHE WOULD OFTEN BE ABLE TO EITHER TURN ON THE SPOT WITHOUT | “IT MEANT SHE WAS MORE INDEPENDENT, AND MAYBE NOT DEPENDENT ON ME.” |
|  | FALLING OR GRAB A BOTTLE FROM THE TABLE WITH FIRMNESS, AND THIS WAS SEEN DURING THE FIRST PHASE, WITH NOTED DECLINE IN THE SECOND PHASE." |  |
| Patient & Parent (quotations provided are from Patient & Parent) | "MORE FLUENT SPEECH" | "COULD TALK MORE WITH HIS MOTHER." |
| Patient & Parent (quotations provided are from Patient & Parent) | "IMPROVEMENT IN SWALLOWING & SPEECH, PATIENT IS MORE "AWAKE", WALKS FASTER & FINE MOTOR IS FASTER" | "SHE IS MORE AWAKE & CAN STAY LONGER AWAKE, SHE CAN TALK EASIER. HOWEVER SHE SWEARS NOW MORE. SHE IS PRETTY MUCH MORE CONTINENT WITH URINE & STOOL. [The changes were] VERY RELEVANT. SHE EXPRESSES FEELINGS NOW ("MOM, I'M COLD./ I'M HUNGRY"), SHE TALKS AND REACTS FASTER, SHE SWALLOWS BETTER." |
| Parent | "HE COULD DRESS UP HIMSELF. HE COULD FOLLOW ORDERS. HE DID NOT GIVE UP PUTTING HIS SHOES ON. HE COULD FOCUS BETTER. HE RESPONDED BETTER. SPEECH WAS BETTER." | "VERY RELEVANT. HE WAS SOCIALLY MORE INTEGRATED." |
| Parent | "MORE ATTENTION, UNDERSTANDS SMALL PROMPTS. WALKS MORE STEADILY, MAYBE FASTER. [I can] SCOLDS A LITTLE LESS." | "VERY MEANINGFUL" |
| Parent | "COGNITIVE IMPROVEMENT, HE REMEMBERES EVERYTHING FAST, MEMORY IMPROVEMENT, WALKING MORE STABLE & FASTER. HE RAISES HIS LEGS BETTER" | "MASSIVE IMPROVEMENT. THEY HAVE STARTED PRACTICE HIS WALKING INDOOR. MASSIVE IMPROVEMENT OF ABSENCES (FROM >5 TO MAX 1 X DAY), SLIGHT IMPROVEMENT OF SWALLOWING. HE CAN FOLLOW BETTER INSTRUCTIONS." |
| Patient | "LESS COUGHING/ NO COUGH WHILE EATING, BETTER SWALLOWING, LESS TIRED/ MORE ENERGETIC. NO MORE FALLS!" | "VERY MEANINGFUL, NO FALLS, NO SWALLOWING PROBLEMS" |
| Parent | WE NOTICED AN IMPROVEMENT IN OUR SON'S LANGUAGE. THE COMMUNICATION WITH OTHERS OUTSIDE OUR FAMILY IMPROVED; OUR SON MADE HIMSELF UNDERSTOOD MORE EASILY." | "THE COMMUNICATION WITH PEOPLE OUTSIDE OUR FAMILY WAS EASIER." |
| Patient | "IMPROVED SIGNIFICANTLY. IMPROVE MOOD, SPEECH, WALKING." | "YES. MADE PATIENT MORE POSITIVE" |
| Patient | "SLIGHT IMPROVEMENT IN OVERALL DAILY ACTIVITIES. HAND DEXTERITY IMPROVE." | "LESS BURDEN ON CARER" |
| Patient | "MY FINE MOTOR SKILLS IMPROVED AND I FELT MORE AWAKE" | "THANKS TO THE IMPROVEMENTS, I CAN TAKE PART IN EVERYDAY LIFE AND ENJOY ME LEISURE ACTIVITIES. THANKS TO MY IMPROVED ALERTNESS / FEELING MORE AWAKE I TOOK PART IN MORE LEISURE ACTIVITIES AND FELT GENERALLY BETTER." |
| Parent | "TAKING PART IN LEISURE ACTIVITIES IS EASIER AND HIS STAMINA IMPROVED." | "THANKS TO THE IMPROVEMENTS, MY SON CAN TAKE PART IN MORE LEISURE ACTIVITIES AND HAS MORE JOY IN LIFE." |
| Parents | "THERE WERE IMPROVEMENTS IN HER FINE MOTOR SKILLS AS WELL AS HER GENERAL MOTOR SKILLS AND BALANCE. THERE WERE SIGNIFICANT IMPROVEMENTS IN OUR DAILY LIFE. REGULAR ACTIVITIES ARE POSSIBLE MORE OFTEN WITHOUT LIMITATIONS." |  |

## Discussion

Previous validation data on the SARA scale has demonstrated construct validity, internal consistency, and interrater reliability, and high reproducibility and responsiveness in patients with inherited Cerebellar Ataxias [2]. This analysis demonstrated the reproducibility, reliability, and responsiveness of the SARA scale in patients with LSDs, and the results of qualitative and quantitative analysis support the SARA scale as a valid measure of neurological status in LSDs that feature central nervous system involvement.

Test-retest data indicate a high degree of consistency between three distinct study cohorts and a high degree of consistency/reliability between visits conducted 2-3 weeks apart (ICC > 0.96), as well as for visits conducted 14-15 weeks apart (ICC > 0.95). The mean change in SARA between assessments in all trials was small indicating that learning effects and other systematic biases are not significant. Notably, the SARA was also demonstrated to be reliable/reproducible in patients <10 years of age. Responsiveness measured as SARA’s ability to classify whether patients had improved or not was above the discrimination threshold for all three Investigator, Caregiver and Patient CGI-I measures (0.82, 0.71 and 0.77 respectively). Notably, the Caregiver CGI-I and Patient CGI-I could be accurately classified with the directional change in SARA (as neither the caregiver or patient are responsible for assessing the SARA scale) and the analysis supports the use of SARA as an endpoint that can detect changes that patients and caregivers consider clinically meaningful.

According to the US Food and Drug Administration, for a clinical endpoint to be meaningful, it should properly reflect or describe how a patient feels, functions, or survives [18]. The high degree of agreement between the SARA scores and the investigator’s, caregiver’s, and patient’s CGI-I, as well as significant improvements in everyday function and quality of life captured in the IB1001-301 exit interviews, support the establishment of a meaningful change threshold of 1-point on the SARA (e.g. a clinically meaningful improvement at -1 point or greater, or a clinically meaningful worsening of +1 point or greater). This was further supported by an analysis of the IB1001-301 exit interviews, where patients/caregivers described how a transition of 1 point or greater reflected the gain or loss of complex functions that were highly relevant to everyday activities, function, and quality of life. That a 1-point change on the SARA is clinically meaningful is consistent with previous literature and the nature of the assessment [2]. The gradation of scoring in the 8 SARA test items was defined to cover the full range of disease severity (from asymptomatic to unable to perform the task in any fashion) and the full spectrum of abilities between these 2 extremes [1]. Thus, each score can be considered to reflect a distinct degree of disease progression and distinct neurological function, so that a 1-point difference is meaningful clinically as observed by the Investigator assessors, and importantly reflects a meaningful difference to a patient’s quality of life.

Our analysis demonstrated that the SARA may be utilized as an outcome assessment in LSDs that feature central nervous system involvement as a wider measurement of neurological function, far beyond the assessment of cerebellar ataxia. Analysis of the exit interviews supports that the SARA scale, when applied and assessed in complex diseases like LSDs that feature a range of heterogenous neurological symptoms, represents a broad assessment of neurological status, namely to signs and symptoms of cortical (understanding of instructions and other cognitive functions, motivation, and planning of movements), basal ganglia, cerebellar, brainstem, pyramidal and extrapyramidal tracts function and dysfunction. The findings from this analysis are supportive of the SARA assessment as a reliable measure of neurological function in patients with LSDs who present with neurological signs and symptoms.

## Data Availability

All authors had full access to all the data in the study and had final responsibility for the decision to submit for publication. The study documents related to the study, and datasets generated and analyzed during the current study are not publicly available. No individual, de-identified participant data will be shared.

## Supplemental Material I – Exit Interview Template

### Phase I

1. Please describe in your own words your (or the patient’s) NPC symptoms before the IB1001-301 trial:
2. Please describe in your own words how your (or the patient’s) NPC symptoms affected your* everyday life before the IB1001-301?

### Phase II

3. Please describe in your own words how your (or the patient’s) experience with NPC symptoms changed during the IB1001-301 clinical trial?
4. Please describe in your own words any improvements (of any magnitude) you observed during the IB1001-301 clinical trial?
5. (If applicable) Please describe in your own words how these improvements (of any magnitude) were relevant/meaningful to you/ your* everyday life?

### Phase III

6. Please describe in your own words any differences (of any magnitude) observed in in your (or the patient’s) NPC symptoms during Period I versus Period II?
7. (If applicable) Please describe in your own words how these differences (of any magnitude) were relevant/meaningful to you/ your* everyday life?

*\*If completed by the caregiver, please describe how these symptoms affected both the caregiver and patient’s everyday life*.

