## Supplementary figures and images for "Assessment of the Reliability, Responsiveness, and Meaningfulness of the Scale for the Assessment and Rating of Ataxia (SARA) for Lysosomal Storage Disorders"

### Figure 1 A & B

**Fig. 1 – Study Schemes from IB1001 Clinical Trials**
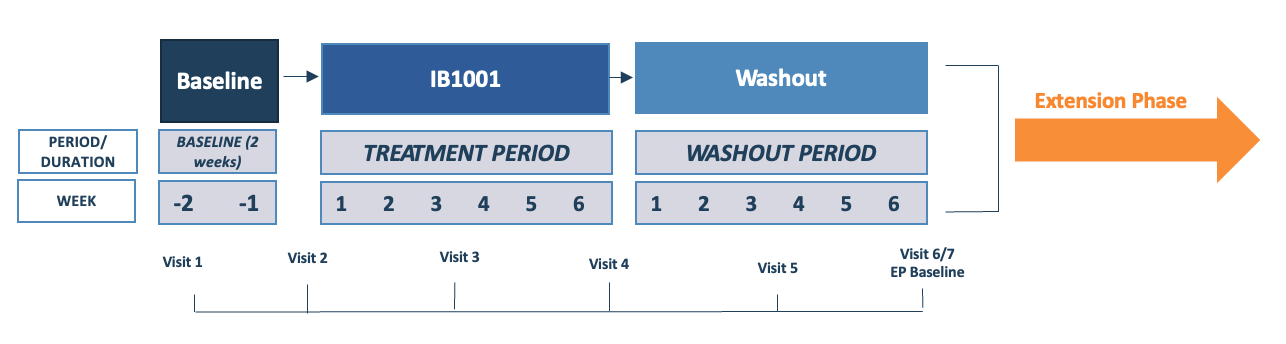

(A) **IB1001-201 / IB1001-202**

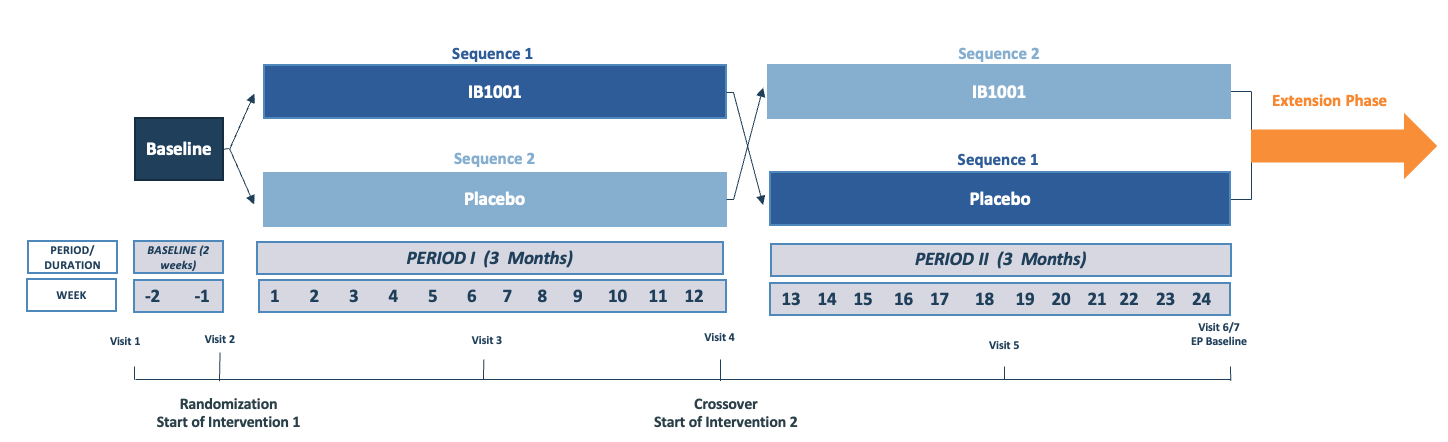

(B) **IB1001-301**
